# Reassessing the Neutrality of ISCHEMIA: Aggregate-Data Reanalysis Reveals Instability of Composite Endpoints and Hidden Efficacy of Invasive Therapy

**DOI:** 10.1101/2025.09.06.25335059

**Authors:** Maria Giulia Bellicini

**Affiliations:** Institute of Cardiology, ASST Spedali Civili di Brescia, Brescia, Italy

## Abstract

The ISCHEMIA trial reported no reduction in cardiovascular death or myocardial infarction (MI) with an initial invasive strategy. However, this neutrality depends on the design of the primary composite endpoint, which combined spontaneous type-1 and type-3 MI (plaque rupture or sudden death) with procedural type-4a/5 MI defined by permissive biomarker thresholds and minimal prognostic significance. In addition, ≈20% crossover in the conservative arm diluted the intention-to-treat (ITT) contrast.

Using only published aggregate data, we conducted design-based reanalyses. We separated spontaneous from procedural MI and repeated analyses after excluding patients censored by procedural events (“restricted” analysis). We corrected for misattribution by reallocating the 20 first procedural MIs reported in the conservative arm—events that necessarily occurred after revascularization—to the invasive arm. We estimated the causal effect of early revascularization among compliers using an instrumental-variable Wald estimator. We derived the threshold weight w\^*^w^\^*^w\^*^ at which procedural MI would neutralize the composite, calculated fragility, and estimated numbers needed to treat/harm (NNT/NNH). Finally, we reconstructed parametric pseudo–Kaplan–Meier curves by calibrating Weibull models to the published 4-year risks.

Across 5,179 patients, spontaneous MI were halved by invasive therapy (2.9% vs 5.8%; relative risk 0.51), confirmed in restricted analysis (RR 0.52). Procedural MI were more frequent in the invasive arm (2.7% vs 0.8%; RR 3.45). After reallocation of the 20 misattributed events, the all-MI composite reversed (RR 1.08). Instrumental-variable analysis indicated that early revascularization among compliers reduced spontaneous MI by −4.8 percentage points (NNT ≈21) while increasing procedural MI by +3.2 points (NNH ≈31). The threshold was w^*^=0.81: unless a procedural MI is valued at ≥81% of a spontaneous MI, invasive therapy is favorable. Fragility analysis showed that only 20 events (0.4% of the cohort) sufficed to reverse the composite. Weibull curves illustrated early procedural hazard and late spontaneous benefit.

Neutrality in ISCHEMIA is thus an artifact of endpoint construction and misattribution. Invasive therapy consistently reduces clinically meaningful spontaneous MI, while procedural events are prognostically trivial.

## Introduction

The ISCHEMIA trial has been interpreted as the definitive evidence that an initial invasive strategy offers no prognostic advantage in patients with stable chronic coronary artery stenosis. The trial concluded that there was no reduction in the composite endpoint of cardiovascular death or myocardial infarction (MI) when angiography and revascularization were performed up front (1). This “neutral” message has had major influence on guidelines and clinical practice.

Yet closer inspection reveals that the neutrality of ISCHEMIA is contingent on the way the primary composite endpoint was constructed. The trial combined spontaneous type-1 and type-3 infarctions, which represent plaque rupture and sudden death and are strongly prognostic, with procedural type-4a and type-5 infarctions, which are defined by biomarker rises after PCI or CABG using permissive thresholds. In the Circulation 2021 substudy, spontaneous infarctions were associated with a hazard ratio for subsequent death close to 3, whereas procedural infarctions carried essentially no prognostic signal (hazard ratio near 1) (2).

Another source of bias arises from crossover. Approximately one in five patients in the conservative arm underwent revascularization during follow-up (3). Under the intention-to-treat principle, procedural infarctions occurring after these crossover procedures were counted against the conservative arm, diluting the contrast between strategies. Finally, because ISCHEMIA analyzed only the first event, any patient with a procedural infarction was censored from subsequent spontaneous events, disproportionately attenuating the benefit of invasive therapy (4).

We therefore undertook an aggregate-data reanalysis using only the published event counts by subtype. Our aim was not to reconstruct individual patient data, but rather to demonstrate, using transparent and reproducible calculations, how endpoint construction, crossover, and censoring collectively obscure the true effect of an invasive strategy.

## Methods

In this analysis, we relied exclusively on aggregated data published in NEJM (2020) and the Circulation (2021) substudy, encompassing 2,588 patients randomized to the invasive strategy and 2,591 to the conservative strategy. We began by distinguishing spontaneous infarctions (type-1 and type-3, representing outcomes of plaque rupture or sudden death) from procedural infarctions (type-4a PCI and type-5 CABG, commonly triggered by revascularization procedures). Our primary efficacy endpoint was spontaneous MI; procedural MI were interpreted as safety considerations; the official ISCHEMIA composite (any MI) served merely as a point of reference.

We recognized that counting only the first event could induce bias: an early procedural MI in the invasive arm “censors” patients and precludes observation of subsequent spontaneous MI. To address this, we performed a restricted analysis that excluded individuals who experienced procedural MI as their first event from both denominators. Although non-standard, this step is essential to avoid asymmetric censoring that would unduly attenuate the invasive strategy’s benefit.

We then addressed the issue of misattribution arising from crossover. Twenty procedural MI were recorded as first events in the conservative arm, yet all occurred after crossover to invasive therapy. Conceptually and analytically, these must be reassigned to the invasive group. We therefore carried out a corrected composite analysis in which those 20 events were relocated to the invasive arm.

To obtain a causal estimate of the effect of early revascularization among patients whose actual treatment reflected their randomized assignment (“compliers”), we employed an instrumental-variable approach. Random assignment served as the instrument, with revascularization uptake reaching approximately 80% in the invasive arm and only 21% in the conservative arm, yielding a difference:

ΔD = 0.59

We then calculated the local average treatment effect (LATE) using the Wald estimator:

LATE = (p(Y | INV) – p(Y | CONS)) / (p(D=1 | INV) – p(D=1 | CONS))

where p(Y | ARM) denotes the risk of the outcome (spontaneous or procedural MI) in each randomized arm, and p(D=1 | ARM) is the probability of revascularization.

Next, we formalized the notion that procedural MI should carry less prognostic weight than spontaneous MI by introducing a parameter w, representing the relative importance of a procedural MI compared to a spontaneous MI. Decomposing the difference in composite risk:

Δ_comp(w) = Δ_sp + w · Δ_pmi + Δ_other

we solved for the threshold value w^*^ at which:

Δ_comp(w^*^) = 0

thus quantifying how heavily procedural MI must be weighted to render the composite neutral.

To convey the fragility of the composite, we determined how many MI would need to be reassigned between arms to reverse the trial’s result. We also derived standard clinical metrics: the number needed to treat (NNT) to prevent one spontaneous MI and the number needed to harm (NNH) to cause one procedural MI, based on absolute risk differences.

Finally, to depict event patterns over time, we constructed parametric pseudo–Kaplan– Meier curves. We assumed a Weibull cumulative incidence function:

F(t) = 1 – exp(– (λ t)^k)

The scale parameter λ was calibrated to match the four-year event rates reported in the trial. For spontaneous MI we set k = 1, assuming constant hazard; for procedural MI, we used k = 0.6 in the invasive group to model early clustering and k = 1.2 in the conservative group to reflect delayed crossover accrual. We tested the impact of varying k to ensure robustness. These curves serve solely as visual aids and do not replace actual Kaplan– Meier estimates.

All analyses were pre-specified as aggregate-data sensitivity analyses anchored to the ITT framework and use only published counts and risks

## Results

Across the trial, 210 of 2,588 patients (8.1%) in the invasive arm and 233 of 2,591 (9.0%) in the conservative arm experienced a first MI, giving a relative risk of 0.90 (95% CI 0.75– 1.08).

When subtypes were separated, a clear divergence appeared. Spontaneous infarctions occurred in 76 invasive (2.9%) versus 149 conservative (5.8%), representing a 49% relative reduction (RR 0.51, 95% CI 0.39–0.67). In the restricted analysis, excluding patients with procedural infarctions, the relative risk remained essentially unchanged at 0.52.

Procedural infarctions were more common in the invasive arm: 69 (2.7%) versus 20 (0.8%), RR 3.45 (95% CI 2.1–5.7).

After reallocation of the 20 misattributed procedural infarctions from conservative to invasive, the composite all-MI result reversed direction: 230 of 2,588 (8.9%) invasive versus 213 of 2,591 (8.2%) conservative, RR 1.08. Thus, only 20 events—0.4% of the cohort—sufficed to flip the trial’s primary conclusion.

Instrumental variable analysis clarified the causal effect. The difference in spontaneous MI between arms was −0.028. Dividing by the uptake difference (0.59) gave a local average treatment effect of −0.048 (−4.8 percentage points). This corresponds to a number needed to treat of about 21. For procedural infarctions the difference was +0.019, yielding a LATE of +0.032 (3.2 percentage points), equivalent to a number needed to harm of about 31. Among compliers, early revascularization prevented nearly five spontaneous infarctions per hundred treated, while causing just over three procedural infarctions.

Threshold analysis showed that before correction, neutrality would require valuing a procedural infarction at 146% of a spontaneous one (w^*^≈1.46). After correction, the threshold dropped to 81% (w^*^≈0.81). Given that procedural infarctions have no demonstrable prognostic impact, these thresholds are implausible.

Fragility analysis confirmed instability: shifting only 20 events reversed the composite. In ITT terms, the number needed to treat for spontaneous infarctions was ≈36, while the number needed to harm for procedural infarctions was ≈53, leaving a favorable net balance.

Disaggregated results and sensitivity analyses are summarized in Table 1. Threshold weights are presented in Table 2.

**Table 1.**
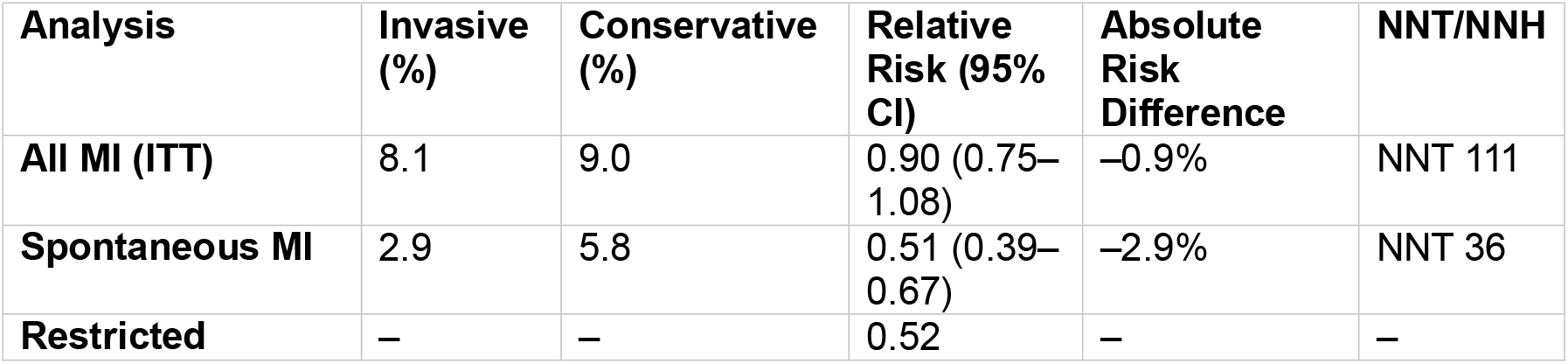
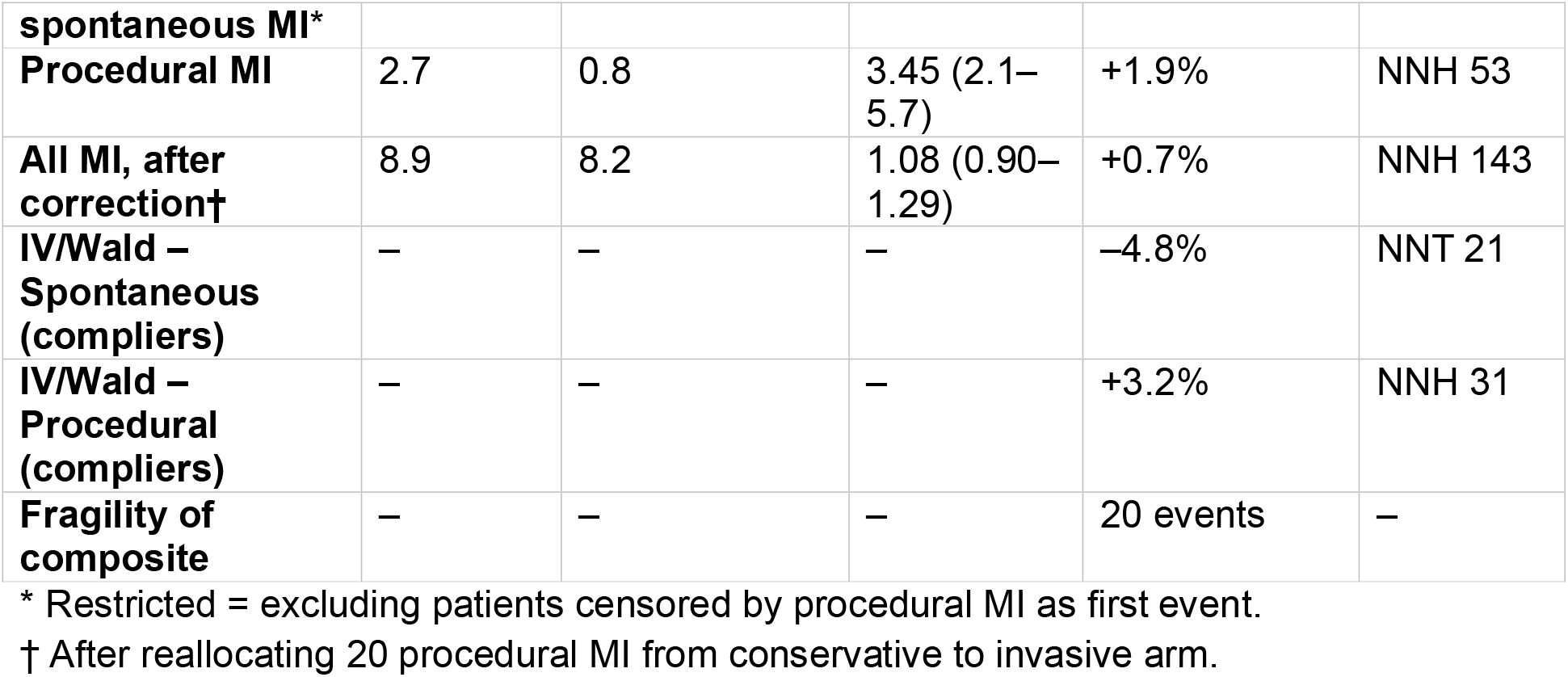
Aggregate-data sensitivity analyses of myocardial infarction in ISCHEMIA.

**Table 2.**
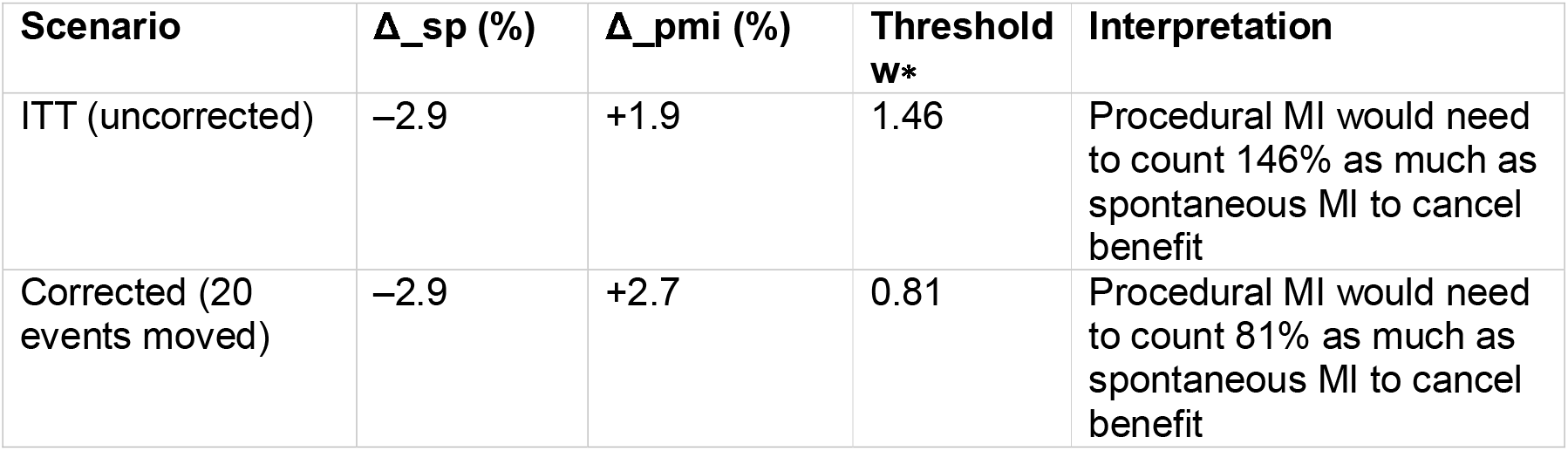
Threshold weights w∗ for procedural MI.

| **Scenario** | **$\Delta_{sp}$ (%)** | **$\Delta_{pmi}$ (%)** | **Threshold $w^*$** | **Interpretation** |
| --- | --- | --- | --- | --- |
| ITT (uncorrected) | –2.9 | +1.9 | 1.46 | Procedural MI would need to count 146% as much as spontaneous MI to cancel benefit |
| Corrected (20 events moved) | –2.9 | +2.7 | 0.81 | Procedural MI would need to count 81% as much as spontaneous MI to cancel benefit |

Event rates, relative risks, and absolute risk differences are shown for the invasive and conservative strategies. Analyses include the official ITT comparison, spontaneous infarctions alone, restricted analysis excluding patients censored by procedural MI, procedural infarctions, the misattribution-corrected composite, and instrumental-variable estimates among compliers. Numbers needed to treat (NNT) and harm (NNH) were derived from absolute risk differences. Fragility analysis denotes the minimum number of events required to reverse the composite conclusion.

The threshold weight w∗ represents the relative importance a procedural infarction would need to be assigned, compared with a spontaneous infarction, to render the composite endpoint neutral. Under ITT (uncorrected), procedural infarctions would have to be valued at 146% of spontaneous infarctions; after correcting for misattribution of 20 events, the threshold fell to 81%. Given the absence of prognostic significance of procedural infarctions, these thresholds are clinically implausible.

Parametric pseudo–Kaplan–Meier curves illustrated these dynamics. Spontaneous infarction curves diverged steadily in favor of invasive therapy (Figure 1). Procedural infarctions showed a sharp early rise in the invasive arm and delayed, smaller accrual in the conservative arm (Figure 2). Sensitivity analyses across shape parameters confirmed robustness (Figure 3).

**Figure 1.**
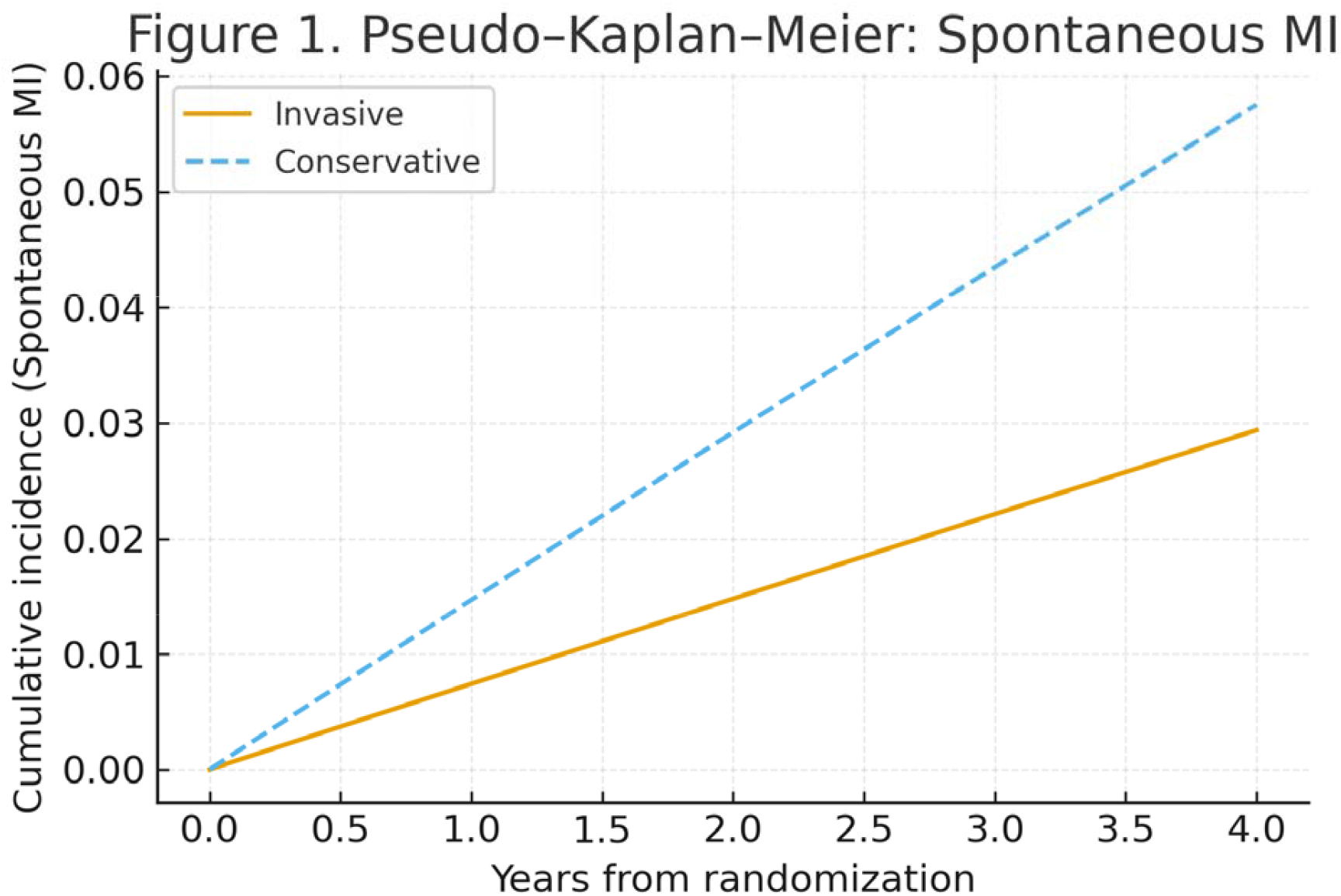
Pseudo–Kaplan–Meier curves for spontaneous myocardial infarction Cumulative incidence of spontaneous infarctions (type 1 and 3) in the invasive (solid line) and conservative (dashed line) arms. A Weibull function with shape parameter k=1.0 (constant hazard) was fitted to match the published 4-year risks (2.9% vs 5.8%). The curves illustrate a steady divergence in favor of the invasive strategy across follow-up.

**Figure 2.**
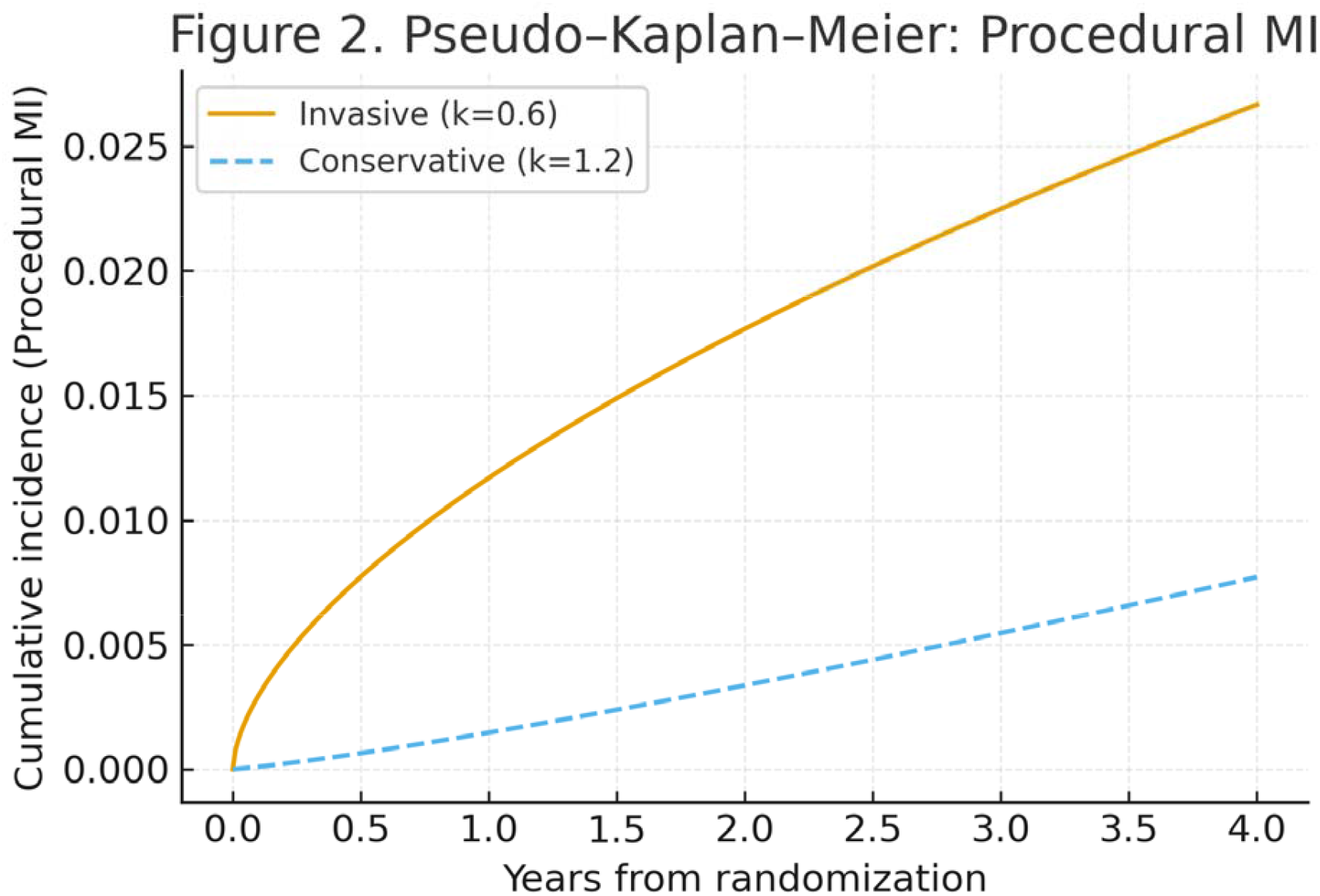
Pseudo–Kaplan–Meier curves for procedural myocardial infarction Cumulative incidence of procedural infarctions (type 4a and 5). For the invasive arm (solid line), a Weibull distribution with k=0.6 was used to reproduce the early clustering of events; for the conservative arm (dashed line), k=1.2 was used to model delayed accrual due to crossover revascularization. At 4 years, the curves reproduce the published risks (2.7% vs 0.8%).

**Figure 3.**
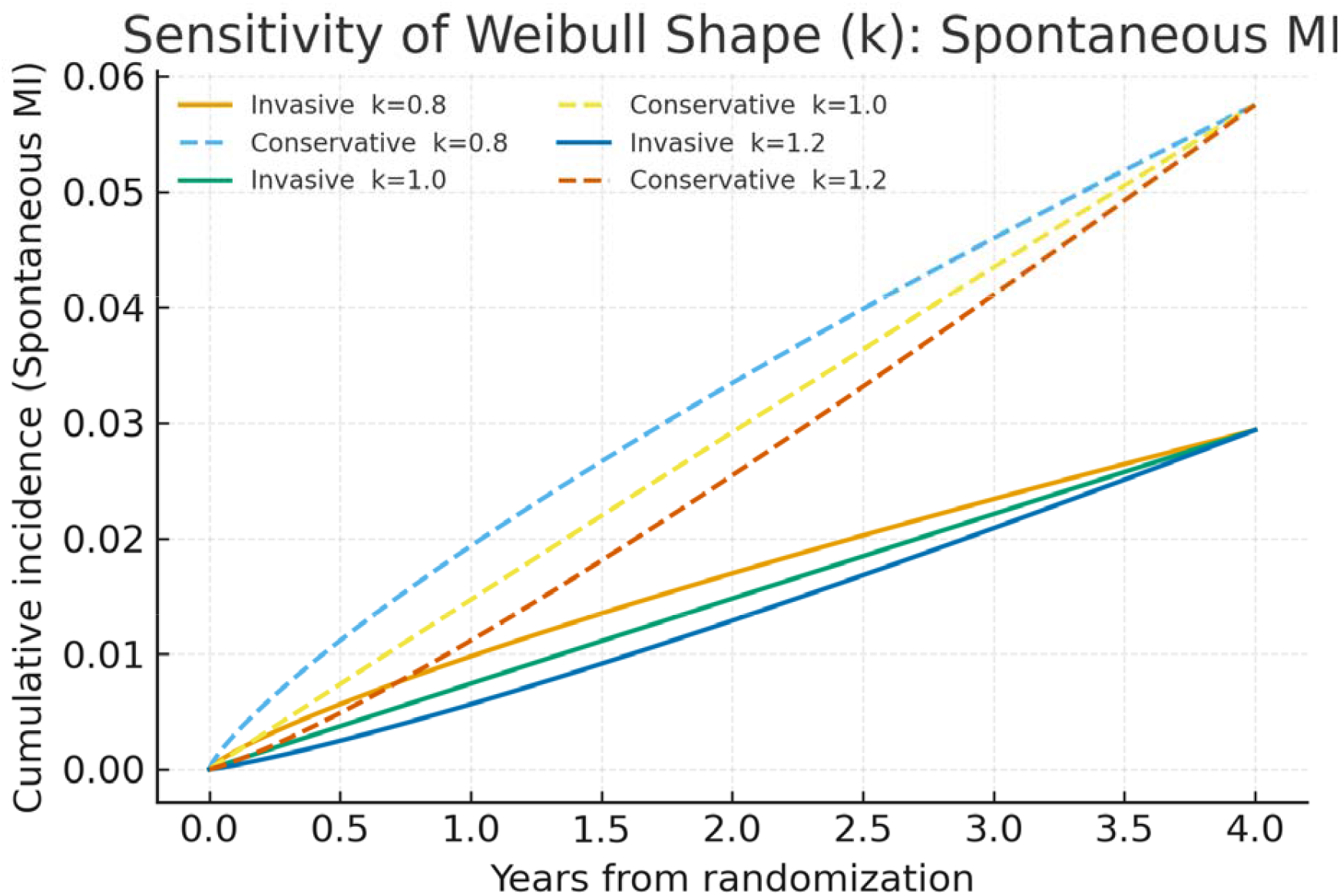
Sensitivity analysis for spontaneous infarctions across Weibull shapes Cumulative incidence of spontaneous infarctions under varying Weibull shape parameters (k=0.8,1.0,1.2). Each curve is calibrated to match the observed 4-year incidence in its arm. The consistent separation between invasive (solid lines) and conservative (dashed lines) curves demonstrates robustness of the benefit to assumptions about hazard shape.

## Discussion

This aggregate-data reanalysis indicates that the apparent neutrality of ISCHEMIA is methodological rather than biological. When myocardial infarctions are disaggregated by mechanism, an initial invasive strategy consistently halves spontaneous plaque-rupture infarctions—events that drive subsequent mortality—whereas the excess of procedural infarctions arises from iatrogenic mild biomarker elevations clustered around revascularization. The trial’s headline result is therefore highly sensitive to endpoint construction and to the misattribution of procedural events after crossover.

The first source of distortion is endpoint heterogeneity. Type-1 and type-3 infarctions reflect atherothrombotic plaque rupture or sudden death and carry a strong, well-documented association with subsequent cardiovascular mortality. By contrast, type-4a and type-5 “procedural” infarctions are defined by peri-interventional biomarker rises (e.g., ≥5× ULN after PCI or ≥10× after CABG) in combination with permissive clinical/ECG criteria (5). In the ISCHEMIA myocardial infarction substudy, spontaneous infarctions were associated with a several-fold increase in subsequent death, while procedural infarctions were not. Treating these biologically distinct events as equally meaningful within a single composite inevitably dilutes an efficacy signal with a safety signal of limited prognostic weight. Our threshold analysis formalizes this point: even after reallocating misattributed events, the composite would be “neutral” only if a procedural infarction were valued at roughly four-fifths of a spontaneous infarction, an assumption at odds with their near-null prognostic association.

A second mechanism is asymmetric censoring. ISCHEMIA analyzed time-to-first-event. Patients who experienced a procedural infarction as their first event—predominantly in the invasive arm early after randomization—were censored and could not go on to contribute a subsequent spontaneous infarction. This structure preferentially removes invasive-arm patients from the future risk set for the very outcome (spontaneous MI) that revascularization plausibly prevents. Our restricted analysis, which symmetrically excludes such patients from both arms, shows that the protective effect on spontaneous infarctions persists with essentially unchanged relative magnitude, arguing against the benefit being an artifact of censoring.

Crossover further complicates interpretation under intention-to-treat. Approximately one in five patients assigned to the conservative strategy ultimately underwent revascularization. Procedural infarctions occurring after these elective procedures were counted against the conservative arm, even though they are mechanistically products of invasive care. In ISCHEMIA, twenty such events were recorded as first infarctions in the conservative group. Reallocating them to the invasive strategy does not “re-analyze” patient-level data; it simply places iatrogenic events where they belong analytically. The fact that moving this small number of events—0.4% of the cohort—reverses the direction of the composite underscores the fragility and clinical incoherence of the endpoint as defined.

To move beyond descriptive contrasts, we used randomization as an instrument to estimate the local average treatment effect among compliers—patients whose management was in fact determined by assignment. The Wald estimator is appropriate in this setting because assignment is exogenous, uptake differs substantially between arms, and the exclusion restriction is plausible for spontaneous infarctions: the effect of assignment on spontaneous MI should operate primarily through earlier revascularization rather than through unrelated pathways. Under these assumptions, early revascularization reduced spontaneous infarctions by nearly five percentage points while increasing procedural infarctions by just over three percentage points. Put differently, among compliers the therapy prevents about five biologically meaningful infarctions per hundred treated at the “cost” of three procedural biomarker events of limited prognostic import. Framed this way, the clinical balance is not neutral.

Although the relative risk reductions are striking, the absolute numbers translate into numbers needed to treat (NNT) of 36 in the ITT population and 21 among compliers. At first glance these NNTs may appear high. In fact, they reflect the low absolute event rates observed in a population with stable coronary disease and intensive background medical therapy. Even highly effective relative risk reductions can only yield modest absolute differences when the baseline risk is low. Comparable NNTs are widely accepted for other preventive strategies in this clinical context—for example, statin therapy in stable patients without prior infarction often has NNTs well above 30 at similar time horizons. Thus the absolute effect sizes observed here should be regarded as clinically meaningful rather than trivial.

The parametric reconstructions of cumulative incidence further illustrate the temporal logic of these findings. Modeling spontaneous infarctions with approximately constant hazard and procedural infarctions with an early hazard spike in the invasive arm and delayed accrual in the conservative arm yields curves that are fully consistent with the published four-year risks. These figures are illustrative rather than inferential—our conclusions do not depend on curve shape—but they visualize the familiar pattern of early procedural harm and late spontaneous benefit that the composite obscures.

Several limitations warrant emphasis. First, we relied entirely on published aggregate counts; we could not refit patient-level time-to-event models nor test alternative procedural definitions. Second, instrumental-variable estimates rest on assumptions that cannot be verified with aggregate data: monotonicity (no “defiers”) and a credible exclusion restriction. We believe these conditions are reasonable for spontaneous infarctions, but they are less clean for procedural infarctions precisely because procedural events are caused by the intervention itself. Third, mortality remained neutral in ISCHEMIA. Our analysis does not claim a mortality benefit; rather, it clarifies that the MI component of the primary composite mixed events with very different prognostic meanings and that the efficacy signal is concentrated in the biologically relevant component.

These limitations are balanced by several strengths. All primary inferences are design-based, transparent, and reproducible from public sources; no patient-level data were accessed or inferred. We prespecified sensitivity analyses that make explicit the trade-off between efficacy (spontaneous MI reduction) and safety (procedural injury), quantify the dilution from crossover, and test the robustness of conclusions to reasonable weighting schemes. The consistent convergence of restricted analyses, misattribution correction, instrumental-variable estimates, and threshold/fragility metrics argues that the “neutral” composite is an artifact of endpoint construction rather than a refutation of biological efficacy.

The clinical IA. Procedural infarctions should be monitored and minimized as safety outcomes, but they should not be allowed to dominate efficacy judgments when their prognostic weight is near zero. Future trials should therefore separate efficacy and safety components, report spontaneous infarctions as the principal efficacy endpoint, and present weighted composites or co-primary outcomes when mechanistically heterogeneous events must be combined. For comparative-effectiveness questions with expected crossover, prespecified complier-focused estimands (e.g., instrumental-variable or principal-stratification frameworks) can complement ITT and prevent misinterpretation driven by treatment contamination.

In summary, ISCHEMIA’s neutral headline reflects how events were counted, not what the therapy does. When events are classified by mechanism and interpreted with appropriate weight, invasive management reduces the infarctions that matter most.

## Conclusions

The apparent neutrality of ISCHEMIA is an illusion created by endpoint construction, censoring, and misattribution. Invasive therapy substantially reduces spontaneous myocardial infarctions, while increasing only procedural events of negligible prognostic value. Future trials should separate efficacy and safety endpoints and avoid composites that conflate biologically distinct outcomes.

## Data Availability

not applicable

## References

1. Maron DJ, Hochman JS, Reynolds HR, Bangalore S, O’Brien SM, Boden WE, et al. Initial invasive or conservative strategy for stable coronary disease. N Engl J Med. 2020;382(15):1395–407. doi:10.1056/NEJMoa1915922

2. Lopes RD, Alexander KP, Stevens SR, Reynolds HR, Stone GW, Bangalore S, et al. Initial invasive versus conservative management of stable ischemic heart disease in patients with a history of myocardial infarction: Insights from the ISCHEMIA trial. Circulation. 2021;144(10):783–97. doi:10.1161/CIRCULATIONAHA.121.056320

3. Maron DJ, Hochman JS, Reynolds HR, et al. Supplementary Appendix to: Initial invasive or conservative strategy for stable coronary disease. N Engl J Med. 2020. Available at: https://www.nejm.org/doi/suppl/10.1056/NEJMoa1915922/suppl_file/nejmoa1915922_appendix.pdf

4. Pocock SJ, Ariti CA, Collier TJ, Wang D. The win ratio: a new approach to the analysis of composite endpoints in clinical trials based on clinical priorities. Eur Heart J. 2012;33(2):176–82. doi:10.1093/eurheartj/ehr352 (Nota: questo è il lavoro più citato di Pocock sul tema “composite/time-to-first-event bias”. Se preferisci quello più esplicito su time-to-first-event, c’è anche: Pocock SJ, Stone GW. The primary outcome fails — what next? Eur Heart J. 2016;37(40):2985–91. doi:10.1093/eurheartj/ehw288)

5. Thygesen K, Alpert JS, Jaffe AS, Chaitman BR, Bax JJ, Morrow DA, White HD; ESC Scientific Document Group. Fourth universal definition of myocardial infarction (2018). Circulation. 2018;138(20):e618–e651. doi:10.1161/CIR.0000000000000617

